# Assessment of Accuracy and Reliability: Validation trial for a Fluorescent Probe-Based Real-Time PCR Assay Kit in Detecting Human Papillomavirus Nucleic Acid

**DOI:** 10.1101/2024.10.29.24316338

**Authors:** Mutamba Barbra, Nyamwiza Alice, Dawit Kassa, Stephen Rulisa

**Affiliations:** University of Rwanda; University Teaching Hospital of Kigali; Eagle Research Center

## Abstract

**Background:** Human Papillomavirus (HPV) infection is a potential risk for cervical cancer, the latter is the fourth leading cause of cancer related death worldwide. Effective testing procedures are particularly important in its prevention and management. More accurate HPV tests replace traditional cytology-based screening, and they are in line with national and international guidelines that recommend primary HPV testing due to its better efficacy. The purpose of the study was to confirm that these Gynius HPV detection kits were equal in efficiency compared with Roche Cobas® HPV assay-based testing, which could better facilitate earlier detection and stratification by risk level for cervical lesions.

**Methods:** The prospective parallel control study recruited 1000 women from the district hospitals of Kibagabaga and Muhima in Rwanda. Participants were tested for HPV DNA tests using both Roche and Gynius kits and then the samples were sent to Rwanda National Reference Laboratory. We analyzed the concordance of two Gynius variants (liquid and lyophilized ) and Roche across for several different HPV subtypes, such as 16, 18 or other high risk types (31,33,35,39,45,51,52,56,58,59,66,68,73,53,82 &26). Statistics like sensitivity, specificity, predictive values and consistency (Kappa) were calculated to compare the concordance of assays.

**Results:** We found that the Gynius HPV solution is exceptionally easy to implement in laboratory settings within low- and middle-income countries (LMICs). It offers effective sampling control through color detection, making it highly suitable for self-sampling procedures. Additionally, the Gynius HPV solution includes an automated extraction system that completes full plate extraction in under 20 minutes.

The Gynius solution is compatible with various qPCR systems, eliminating the need for costly qPCR machine repurchases. Using lyophilized reagents, which can be stored and transported at room temperature, it bypasses the need for a cold chain and refrigeration.

The Gynius solution also has a high sample processing capacity. For instance, using the BioRad CFX96, it can produce up to 1,200 results per day per machine; with the BioRad CFX384, it can generate up to 4,000 results per day—making it ideal for large-scale screening.

In a comparison of 987 samples, high-risk HPV detection rates were consistent across all three tests: Roche (20%), Gynius liquid (23%), and Gynius lyophilized (21%). Concordance analysis showed high agreement rates, with both Gynius kits achieving over 80% compatibility with Roche for detecting HPV 16, 18, and other virus subtypes.

**Conclusion:** This study provides compelling evidence for the efficacy of the Gynius HPV kits (both lyophilized and liquid formulations), which demonstrated high concordance rates with established assays for detecting high-risk HPV genotypes, particularly HPV 16 and 18. Engineered specifically for low- and middle-income countries (LMICs), the Gynius HPV kits integrate innovative sampling and detection technologies optimized for resource-limited settings, facilitating seamless implementation. These findings indicate that Gynius HPV kits present a viable and scalable solution for cervical cancer screening programs, with the potential to significantly enhance early detection capabilities and broaden preventive care accessibility.

## Background

Cervical cancer is a significant global health burden, ranking as the fourth most diagnosed cancer and the fourth leading cause of cancer-related deaths with an estimated 604,000 new cases and 342,000 deaths worldwide in 2020 (1). Its prevalence and mortality underscore the urgency of effective screening and early detection strategies (2). The Human Papillomavirus (HPV) is responsible for most cancers and is thought to be responsible for more than 90% of anal and cervical cancers, about 70% of vaginal and vulvar cancers, and 60% of penile cancers (3). HPV (human papillomavirus) is one of the most common sexually transmitted infections (4). Human Papillomavirus (HPV) is the most prevalent sexually transmitted virus and most attributable to cervical cancer (6,7), it (HPV) has around 100 types of HPV, with different variations in their genetic and oncogenic potential (8).

Traditional cytology-based screening method has been an important cornerstone in programs for the detection of preinvasive cancer. However, the development of Human Papillomavirus (HPV) DNA tests has deftly proven that they are more effective than cytology-based screening in identifying abnormal cells of the uterine cervix (9). Several national and international guidelines now advocate transitioning from cytology to HPV-based primary screening owing to its superior performance and effectiveness in identifying high-risk HPV infections that lead to cervical cancer (4,10).

The adoption of HPV-based primary screening method detects the presence of various high-risk HPV strains (especially types 16 and 18), these DNA tests allow earlier identification of people at risk for developing cervical abnormalities or cancer. In addition, the transition to HPV-based screening is anticipated to significantly reduce the burden of cervical cancer by enabling more accurate and timely detection, subsequently facilitating prompt intervention and treatment (11).

The newly introduced HPV DNA tests for primary screening must go through rigorous steps of validation to gain clinical acceptance, reliability and accuracy (12). This validation process is crucial for establishing the credibility of HPV DNA tests, ensuring their reliability and feasibility in accurately identifying individuals at risk and differentiating between those who require further diagnostic evaluation and those who do not (13).

The primers and fluorescent probes of this kit are specifically designed in the L1 loci of the HPV genome, and the fluorescent probes are labeled with FAM, HEX/VIC, and ROX fluorescent dyes, respectively. Polymerase chain reaction (PCR) combined with Taqman fluorescent probe technique will be used to detect nucleic acids of 18 HPV genotypes in women’s cervical shedding cells. And this kit types HPV16 and HPV18, but does not specifically type the other 16 HPV genotypes. The primer probe is designed according to the human HAPB gene sequence as the internal standard of the kit, and the internal standard probe is labeled with Cy5 fluorophore. The anti-pollution components UDG enzyme and dUTP are also added to this kit. During PCR amplification, DNA polymerase infiltrates dUTP into the PCR product, and UDG enzyme cleaves the N-glycosyl bond between uracil base and sugar phosphate backbone to break the DNA strand containing dU, thus eliminating the contamination caused by PCR product and ensuring the specificity and accuracy of amplification result. This kit contains 5 components: Buffer, Primer/Probes, Enzyme Mixture, Positive Control, Negative Control. The lyophilized reagent contains PCR Mix tubes, Positive control, Negative control, nuclease free water, TE buffer and 8 strip caps.

Furthermore, the efficiency of this investigational kit is underscored by its rapid turnaround time. The entire testing process, from sample analysis to obtaining results, is completed within a remarkably short duration of around 90 minutes. This rapid turnaround time is crucial in clinical settings, as it allows for prompt decision-making regarding further diagnostic evaluation or treatment strategies, thereby facilitating timely interventions. We also found the Gynius HPV kits are designed to the LMICs with certain uniqueness: 1, the sampling kit has color detection for sufficient sampling, 2, automated extraction system to avoid manual operation with high speed, 3, open reagent to most of the qPCR machines, 4, the lyophilized reagent is able to be transported and used under room temperature, 5, high capacity for large scale screening session, 6, low consumables cost, such as pipetting tips, PCR plates, etc.

The aim of this study was to verify the Human Papillomavirus Nucleic Acid Amplification Test Kit (Fluorescent Probe-Based Real-Time PCR Assay). Its relative performance was measured against the widely used “Cobas® HPV” by Roche, which is considered a strong reference test (15). Validating this type of testing kits is likely to have a huge impact on cancer burden because it will help in early detection, as well as subsequent intervention.

## Methods

### Study Design

This study used a prospective parallel control validation design. It compared clinical samples testing using the investigational HPV DNA detection kit (Fluorescent Probe-Based Real-Time PCR Assay) with contrast reagents (Cobas® HPV” by Roche). The primary aim for this design was to assess the effectiveness and applicability of the investigational kit in detecting HPV-related diseases, particularly cervical cancer. In cases of inconsistent test results between the assessment and contrast kits, advanced molecular techniques like next-generation sequencing or genotyping are served for third-party verification. This systematic approach aimed to comprehensively validate the performance of the investigational kit, ensuring its reliability and accuracy for clinical use in detecting HPV-related diseases.

### Participants

One thousand participants from two representative study sites (Kibagabaga and Muhima Hospitals both located in Kigali city were selected based on population differences, epidemiological background, and characteristics of pathogenic microorganisms.

The criteria for inclusion in the study encompassed patients with various clinical presentations, from normal cervical smears to abnormal epithelial cells and direct sequential specimens of CIN or different stages of carcinoma. Eligible subjects needed to submit samples of a certain volume (1mL for Gynius selected extraction reagent and 20 mL for the contrast.

In contrast, exclusion criteria consisted of individuals who practiced vaginal douching or took intravaginal medications (such as antifungal and contraceptive creams) for three days before sample collection. Those had intercourse recently or cervical smears were done with acetic acid or iodine within 24 hours before collection, pregnant women and those in the eighth week after childbirth and those having a history of total hysterectomy. Women that come for a regular cervical cancer screening or for routine obstetric and gynecologic consultations and meeting the inclusion criteria will be enrolled in the study.

Sample size was calculated based on an expected positive/negative coincidence rate of 90%, resulting in an enrollment of at least 552 cases to cover various HPV types and characteristics of enrolled subjects. The total sample reached 1000 which increased the robustness and generalizability of the results.

### Testing methods

This is a multicenter prospective clinical validation study. In this study, three different samples were collected from the same subject that met the inclusion criteria was used to test samples collected from the same subject that met the inclusion criteria using the test and comparison reagents. If the HPV nucleic acid test results of the assessment reagent and the contrast reagent are inconsistent, genotyping is used as the third-party confirmation method.

Patient Volunteers were sampled separately using the Roche and Gynius sampling kits. The Roche samples underwent analysis via the Roche Cobas 4800 system. Gynius samples underwent extraction using the Automated Nucleic Acid Extraction System (Mole 32M) with Mole Nucleic Acid Extraction kit, and subsequently analyzed using the Biorad CFX96 PCR system with two separate reagents.

The two Gynius kits include:

1. Gynius Human Papillomavirus Virus Nucleic Acid Amplification test Kit – liquid form (96Tests/box).
2. Gynius Human Papillomavirus Virus Nucleic Acid Amplification test Kit – lyophilized form (96Tests/box).

Both results were performed by trained laboratory scientist. from the Rwanda national laboratory. Both test were compared and whenever there was a discrepant samples between Roche and Gynius, HPV gene sequencing method was used as the third-party confirmation method.

We used cobas® HPV as reference test (16). This assay is a qualitative in vitro test for Human Papillomavirus (HPV) that has been specially designed to detect the virus on cervical samples taken by clinicians using an endocervical brush, spatula, or broom. For subsequent analysis, these collected samples were preserved in Thin Prep® Pap Test™ PreservCyt®

Solution. Several factors influenced the choice of Roche Cobas® HPV test as reference standard. This test demonstrated a higher level of sensitivity and specificity in detecting various HPV genotypes compared to other available assays (15). In addition, the Roche Cobas® HPV test has received approval from the Food and Drug Administration (FDA) and undergone clinical validation. It is capable of providing individualized results for specific HPV (16,17) making it an excellent tool against which we could compare our investigational kit.

### Estimating diagnostic accuracy and data analysis

Evaluation indicators for parallel detection included positive/negative/total coincidence rates, their 95% confidence intervals, and the Kappa consistency test. Samples with inconsistent results underwent confirmation using the sequencing or genotyping method.

Several evaluation criteria were used to evaluate the investigational reagents and contrast methods, such as the test’s ability to correctly identify individual with high risk HPV (sensitivity) and those without high risk HPV (specificity). Also, the Kappa consistency test (Kappa value) was used to ensure there is agreement between the tests. In each case an assessment of a 95 % confidence interval was made. According to Kappa values, the results of tests were consistent when they reached at least 0.75. Only subject with both reference and investigational test were considered valid and analyzed. This study followed the Standards for Reporting of Diagnostic Accuracy Studies (STARD) (18).

### Ethical Considerations

The study adhered to ethical standards, and before proceeding obtained approval from the Rwanda National Ethics Committee. All participants gave informed consent before undergoing any study-related procedures. This process included a thorough explanation to participants of the study’s purpose, procedures and possible benefits & risk. The risks involved in the study were explained to them. Participants in the study understood completely the nature of their involvement and agreed to participate voluntarily.

Furthermore, after the results of the study were obtained, they were communicated back to these participants. In cases where individuals tested positive in the screening, and such patients needed further evaluation or medical interventions/treatment, they were immediately referred to Kigali’s University Teaching Hospital (CHUK) for a further assessment and follow up.

## RESULTS

A total of 1000 women who met the inclusion criteria were enrolled in the study and underwent cervical cancer screening using HPV DNA testing Using Roche, Gynius kits at Kibagabaga and Muhima district hospitals in Rwanda. Most participants were aged between 25-44 years, with a mean age of 36.6 years (ranging from 15 to 87 years) (table1).

**Table 1:** Age category of the participants.

| Age category | Number | Percentage |
| --- | --- | --- |
| 15-24 | 105 | 10.5 |
| **25-34** | 365 | 36.5 |
| **35-44** | 333 | 33.3 |
| **45-54** | 135 | 13.5 |
| **55-64** | 48 | 4.8 |
| **65-74** | 11 | 1.1 |
| **75-84** | 2 | 0.2 |
| **85-94** | 1 | 0.1 |
| **Total** | **1000** | **100%** |

Out of 1000 women selected for initial screening, 987 were considered valid and included in analysis. Among these participants, the Roche assay detected positive results for high-risk HPV in 20.7 %), individuals (205) while Gynius liquid kit and lyophilized kits both produced positive rates of prevalence at 23.7 % (233) and 21 % (207) respectively (table 2).

**Table 2:**
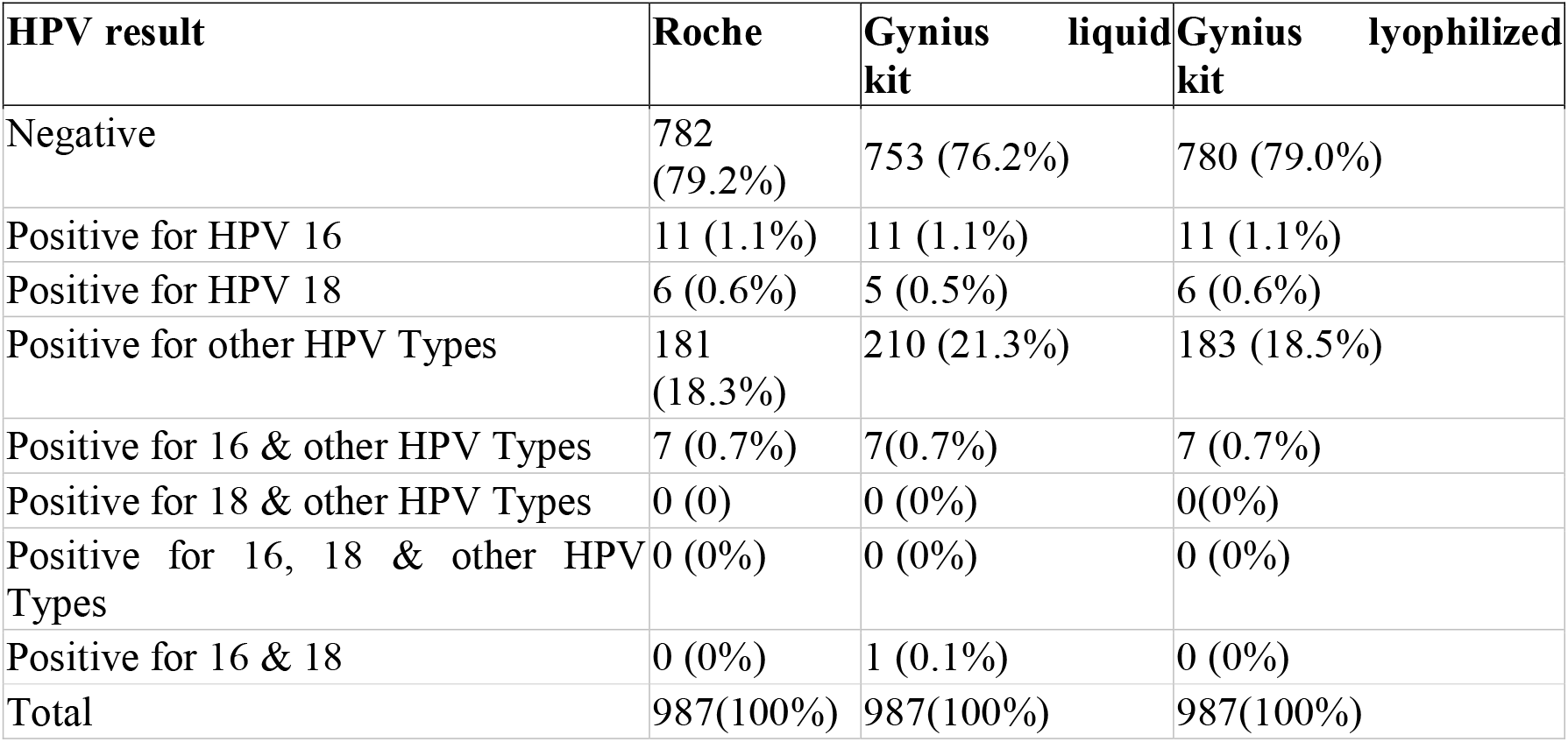
HPV result of women who underwent cervical cancer screening using three HPV DNA assays (Roche and two Gynius kits).

### Concordance analysis

The concordance analysis was performed between Roche and 2 Gynius kits (Gynius liquid and Gynius lyophilized) across 3 HPV categories (HPV 16 subtype, HPV 18 subtype and other HPV subtypes).This was conducted to assess agreement between the results obtained from the Roche kit and two Gynius kits (Gynius liquid and Gynius lyophilized) across three specific HPV categories: HPV 16 subtype, HPV 18 subtype and other HPV types.

### HPV 16 Subtype

These was a 100 % concordance rate between the Roche kit and both the Gynius Liquid Kit and the Gynius Lyophilized Kit. This perfect agreement is also indicated by a Kappa value of 1.000. The sensitivity, specificity, positive predictive value, negative predictive value, and accuracy were all at 100 % (table 3 and 4).

**Table 3:**
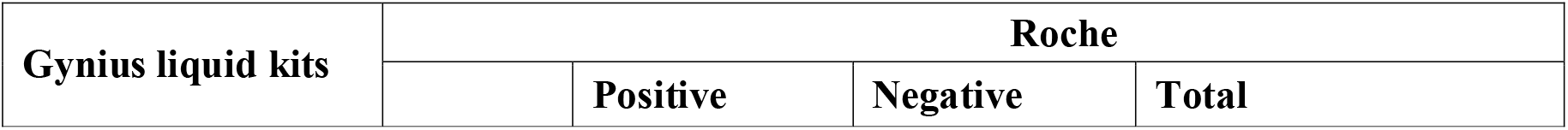
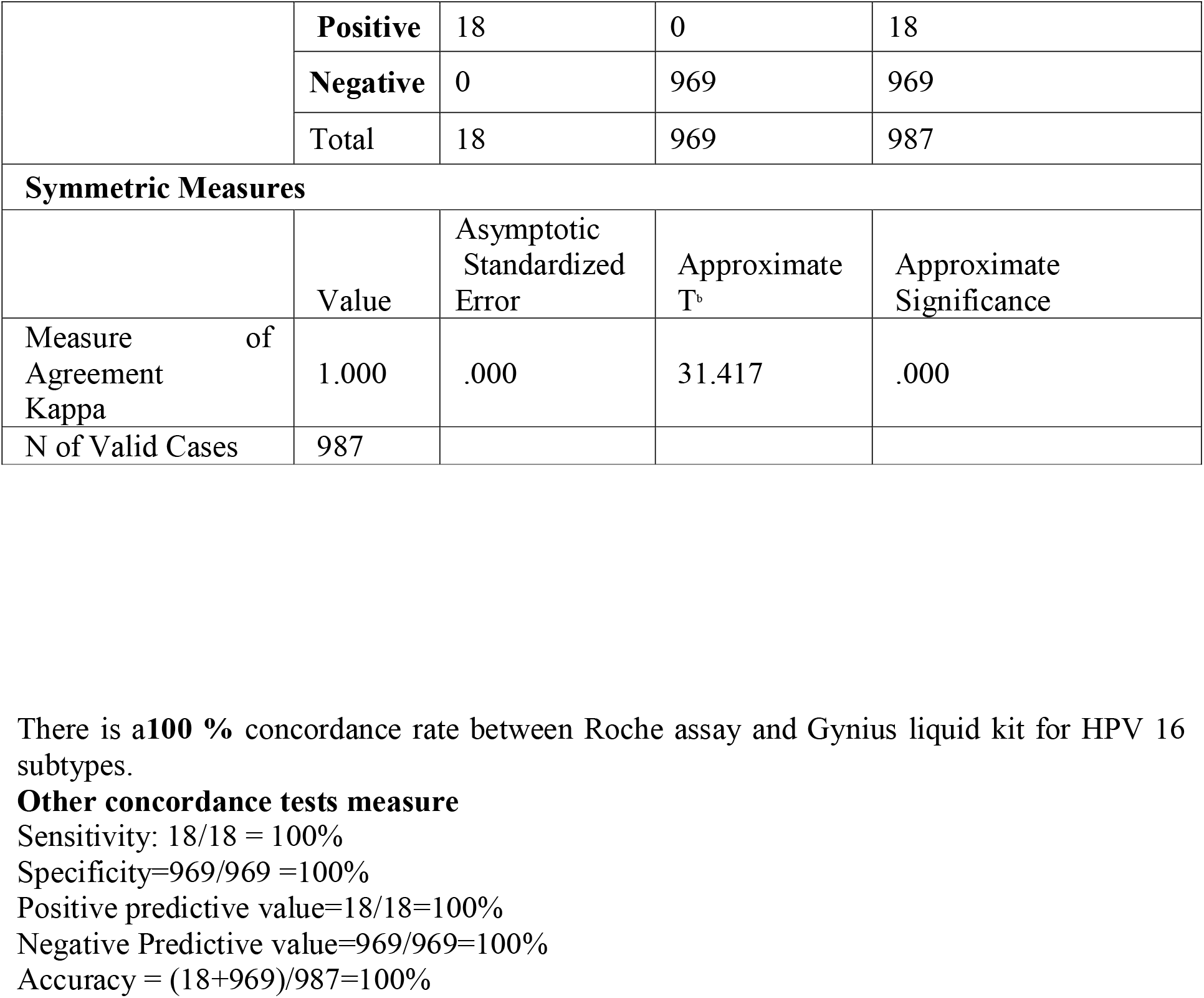
Concordance between Roche and Gynius liquid kits for HPV 16 subtype.

**Table 4:**
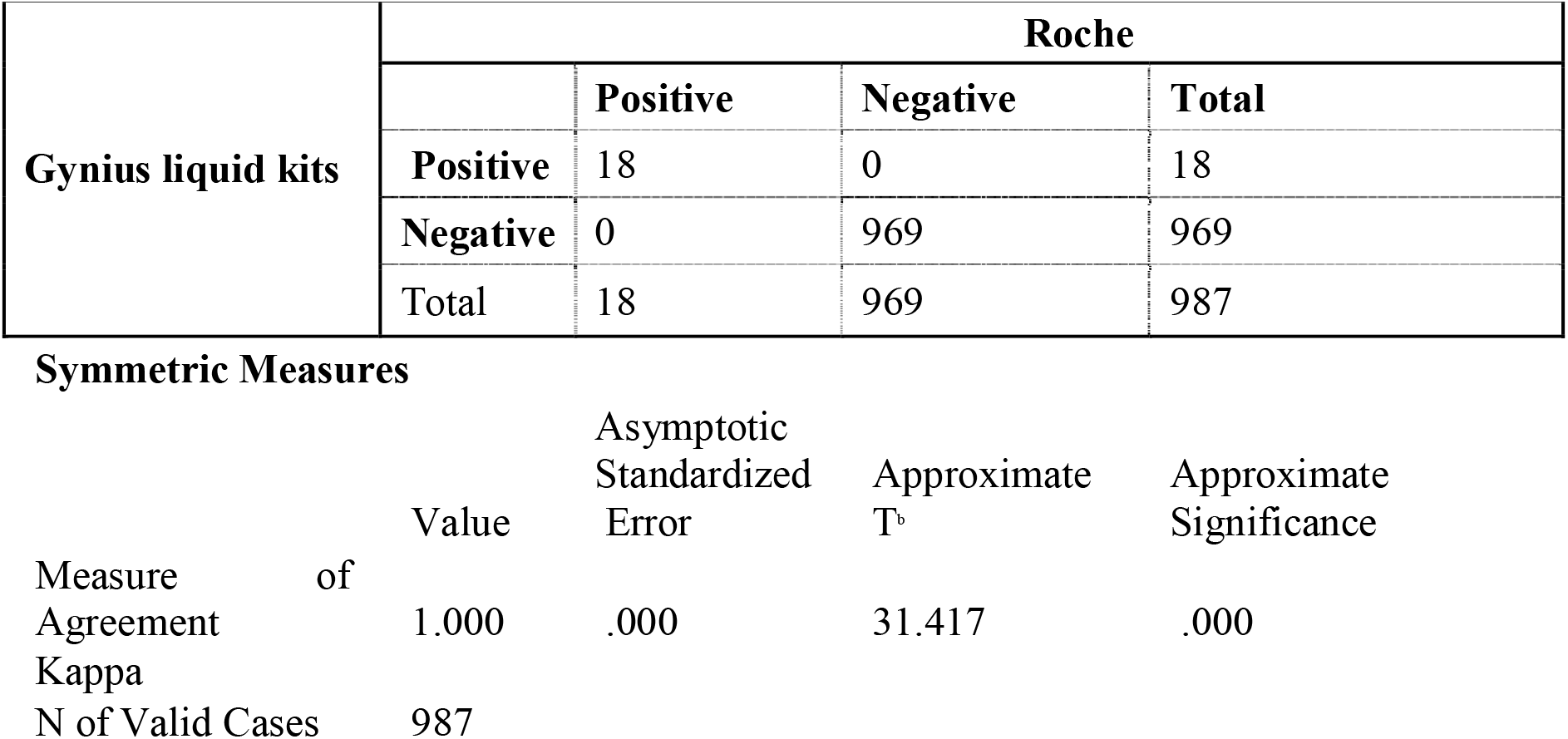
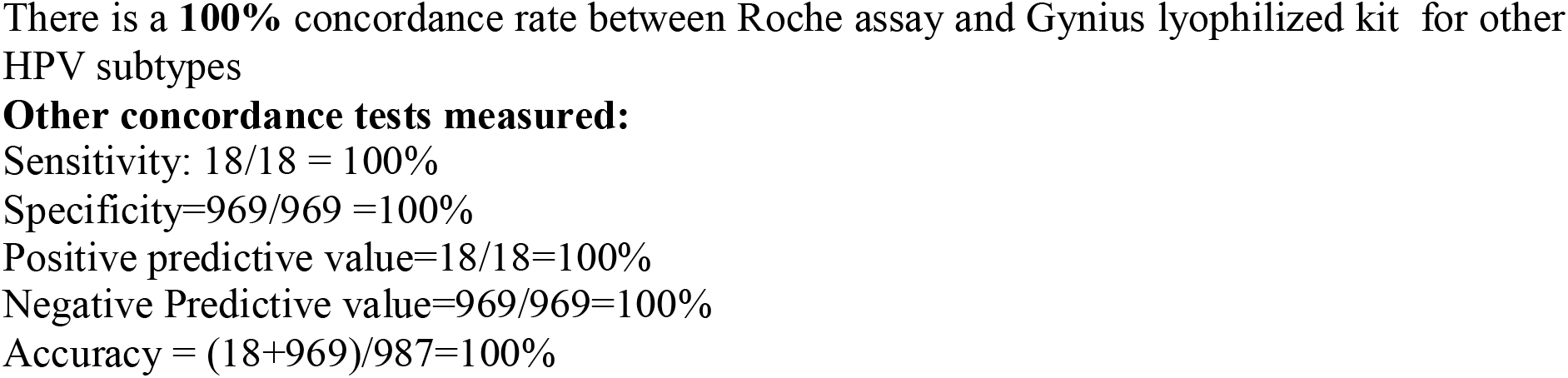
Concordance between Roche and Gynius lyophilized kit for HPV 16 subtype.

### HPV 18 subtype

Roche and the Gynius Liquid Kit agreed at 83.2 % rate (kappa = 0.832), for HPV 18. This points to a high degree of concordance between the two tests. The sensitivity of the Gynius Liquid Kit was thus 83 % and specificity was 99.8 while the positive predictive value (83%), negative predictive value (99.8%), and accuracy (99.7) were all high (table 5).

**Table 5:**
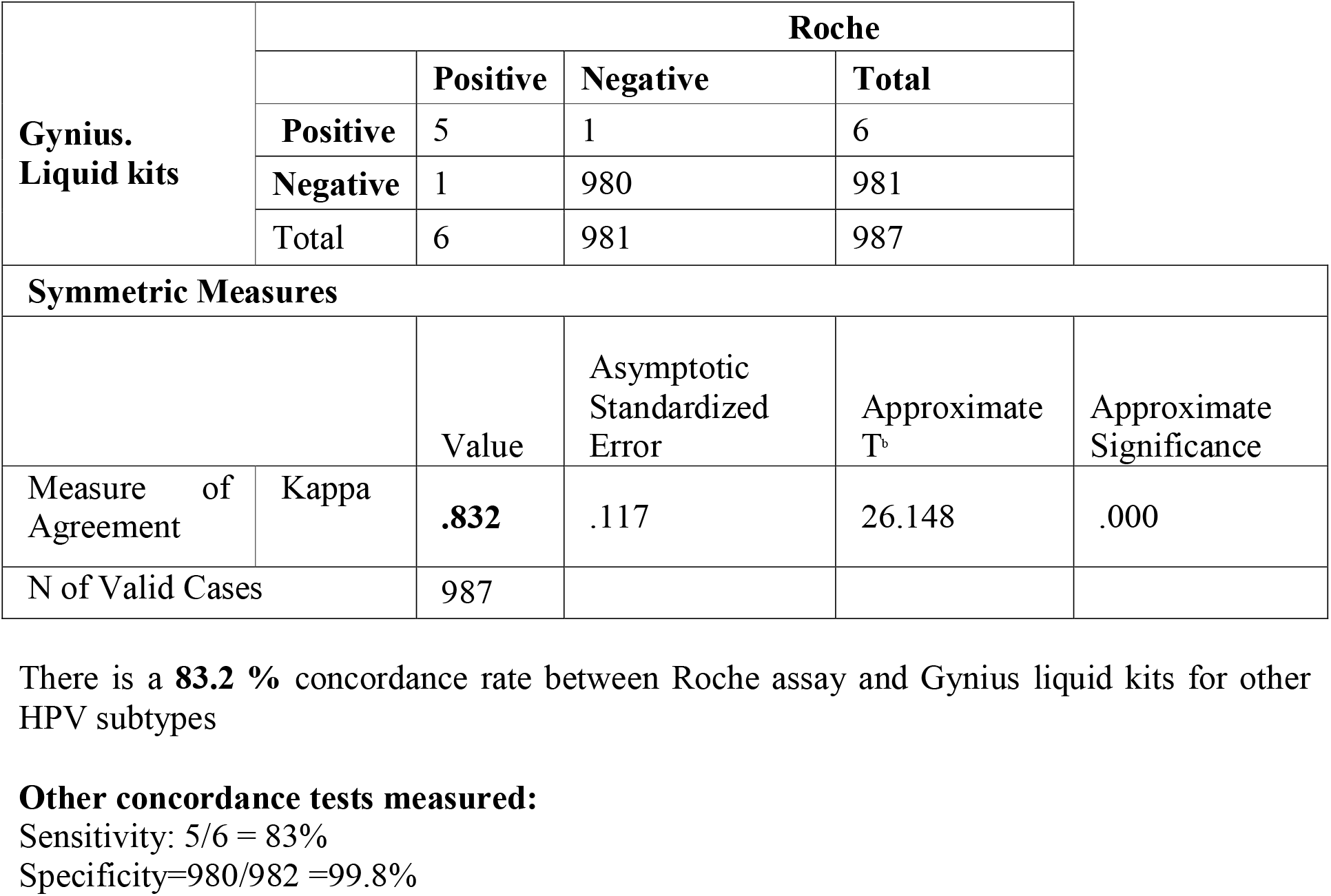
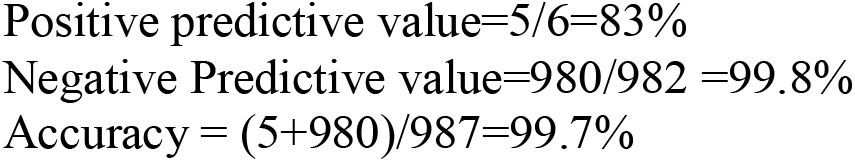
Concordance between Roche and Gynius liquid kits for HPV 18 subtype.

Furthermore, the comparison between Roche and Gynius Lyophilized Kit for HPV 18 subtype (table 6) yielded a match rate of 100 % (Kappa = l.0). This represents full conformity between the two kits. This was the highest sensitivity (100 %), specificity (100 %), positive predictive value (100 %), negative predictive value (100 %), and accuracy (100 %). There was an overall high agreement between Roche and both Gynius kits.

**Table 6:** Concordance between Roche and Gynius lyophilized kit positivity for HPV 18 subtype.

| Gynius liquid kits | Roche |  |  |  |
| --- | --- | --- | --- | --- |
|  |  | Positive | Negative | Total |
|  | Positive | 6 | 0 | 6 |
|  | Negative | 0 | 981 | 981 |
|  | Total | 6 | 981 | 987 |

| Symmetric measures |  |  |  |  |  |
| --- | --- | --- | --- | --- | --- |
|  |  | Value | Asymptotic Standardized Error | Approximate T<sup>b</sup> | Approximate Significance |
| Measure of Agreement | Kappa | **1.000** | .000 | 31.417 | .000 |
| N of Valid Cases |  | 987 |  |  |  |

There is a **100 %** concordance rate between Roche assay and Gynius lyophilized for other HPV subtypes

**Table 7:**
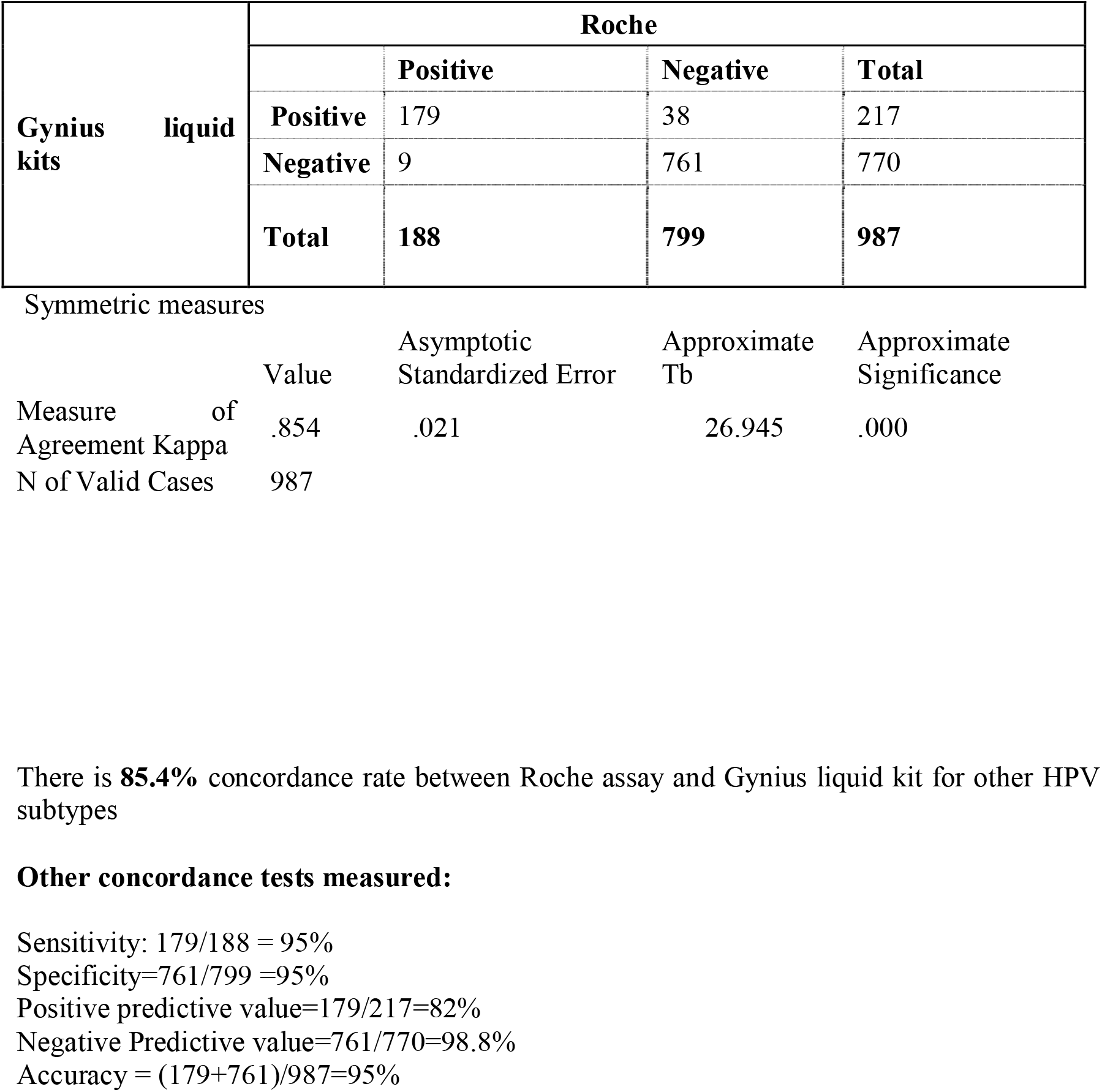
Concordance between Roche and Gynius liquid kits for other HPV subtypes.

**Table 8:** Concordance between Roche and Gynius lyophilized kit for other HPV subtypes.

| Gynius<br>liquid kits |  | Roche |  |  |
| --- | --- | --- | --- | --- |
|  |  | Positive | Negative | Total |
|  | Positive | 167 | 23 | 190 |
|  | Negative | 21 | 776 | 797 |
|  | Total | 188 | 799 | 987 |

Symmetric measures
|  | Value | Asymptotic<br>Standardized<br>Error | Approximate<br>Tb | Approximate<br>Significance |
| --- | --- | --- | --- | --- |
| Measure of Kappa Agreement | .856 | .021 | 26.894 | .000 |
| No of Valid Cases | 987 |  |  |  |

There is a **85.6%** concordance rate between Roche assay and Gynius lyophilized kit for other HPV subtypes

### Other HPV subtypes

As detailed in table 3, there was a relative a strong agreement between HPV subtypes.

- **Roche vs. Gynius Liquid Kit** There was 85.4% agreement between the reference test (Roche) and Gynius Liquid Kit (Kappa value of .854). Sensitivity, specificity, positive predictive value, negative predictive value, and accuracy were 95%, 95%, 82%, 98.8%, and 95%, respectively.
- **Roche vs. Gynius Lyophilized Kit** There was 85.6% agreement between the reference test (Roche) and Gynius Liquid Kit (Kappa value of .856). Sensitivity, specificity, positive predictive value, negative predictive value, and accuracy were 88.8%, 97%, 87.8%, 97%, and 95.5%, respectively.

During validation, several unique features of the Gynius HPV kits were highlighted, underscoring their suitability for low- and middle-income countries (LMICs). The kits include a colorimetric sampling confirmation that ensures sample adequacy, especially useful in self-sampling procedures. An automated high-speed extraction system minimizes manual operation, enhancing throughput and reliability. Unlike other systems, the Gynius kits are compatible with a wide range of qPCR platforms, reducing the need for costly, specialized equipment. With lyophilized reagents that can be stored and transported at room temperature, the kits eliminate cold chain requirements, further simplifying logistics in resource-limited settings. Cost efficiency is particularly high; each sample requires only one 1000 µL pipette tip, one 5 µL tip, half of a 200 µL tip, and does not require a PCR plate. Together, these characteristics make the Gynius HPV kits both practical and economical for large-scale screening programs.

## Discussion

This study evaluated the performance and concordance of the Fluorescent Probe-Based Real-Time PCR Assay Kit (Gynius) against the established Roche assay in detecting HPV nucleic acid. Our objective was to determine whether the Gynius kits could effectively serve as alternatives or complementary tools to the Roche system in HPV detection and genotyping, particularly within the context of low- and middle-income countries (LMICs).

Both Roche and Gynius kits were used to screen for high-risk HPV, with the Gynius kits available in both liquid and lyophilized formats. Results demonstrated high concordance between the two systems, with the Gynius kits detecting HPV positivity at rates comparable to the Roche assay. Notably, concordance was especially strong for HPV 16, where both the liquid and lyophilized Gynius kits showed complete agreement with the Roche assay.

The Gynius kits exhibited several unique features that underscore their suitability for LMIC settings. The colorimetric sampling confirmation in the Gynius kits allowed for enhanced sampling reliability, particularly beneficial in self-collection contexts. The automated, high-speed extraction process reduced manual handling, increasing throughput and minimizing error. Additionally, the Gynius kits’ compatibility with a range of qPCR systems provides adaptability in settings that may lack specialized equipment, lowering the financial barrier to HPV testing. The lyophilized reagents’ room-temperature stability further facilitates their deployment in resource-limited environments by removing cold-chain requirements. Finally, the Gynius kits’ high throughput capacity enables large-scale screening, and the reduced need for consumables such as pipette tips and PCR plates significantly lowers per-test costs.

While this study presents encouraging evidence for the Gynius kits, several limitations should be noted. Selection bias may have been introduced due to the specific hospital-based sample population and the convenience sampling method used. Additionally, reliance on a single testing round without longitudinal follow-up may limit understanding of HPV dynamics over time. While strong concordance rates were observed for key HPV subtypes, statistical uncertainty remains in subtype categories with smaller sample sizes. The generalizability of these findings may also be limited to the studied demographic and healthcare setting, and caution should be exercised when extending these results to broader populations, considering regional HPV prevalence, genetic variability, and healthcare infrastructure.

Overall, despite these limitations, the Gynius kits demonstrated consistent performance with the Roche assay across various high-risk HPV subtypes (including HPV 16, HPV 18, and others), achieving concordance rates above 80%. These findings support the Gynius kits as viable and efficient options for HPV screening, particularly suited for cervical cancer prevention initiatives in resource-constrained settings.

## Conclusion

The results of this study demonstrate strong concordance between the Roche kit and the Gynius HPV kits across various HPV subtypes, with especially high agreement for HPV 16, where both the Gynius liquid and lyophilized kits achieved perfect concordance with the Roche assay. Furthermore, the Gynius kits demonstrated an advantage by detecting a wider range of high-risk HPV subtypes, enhancing their clinical applicability for comprehensive screening.

In addition to high diagnostic accuracy, the Gynius kits incorporate several unique features tailored to low- and middle-income countries (LMICs). With colorimetric sampling confirmation, these kits ensure reliable sample adequacy, which is particularly beneficial for self-sampling. The automated high-speed extraction system reduces the need for manual handling, increasing throughput while minimizing errors. Their compatibility with multiple qPCR systems also makes the Gynius kits adaptable across various lab settings, eliminating the need for specialized, costly equipment. The use of lyophilized reagents, stable at room temperature, removes cold chain requirements, making them ideal for resource-limited environments. Finally, the high throughput capacity and low consumable cost—requiring minimal pipette tips, PCR plates, and other materials—support cost-efficient, large-scale screening programs.

These distinctive advantages underscore the potential of Gynius HPV kits as viable, scalable options for HPV screening in LMICs, promoting broader accessibility and enhanced cervical cancer prevention efforts.

## Data Availability

data is available when requested

